# Exploring the Feasibility of Using Real-World Data from a Large Clinical Data Research Network to Simulate Clinical Trials of Alzheimer’s Disease

**DOI:** 10.1101/2020.06.03.20121491

**Authors:** Zhaoyi Chen, Hansi Zhang, Yi Guo, Thomas J George, Mattia Prosperi, William R Hogan, Zhe He, Elizabeth A Shenkman, Fei Wang, Jiang Bian

**Author notes:** Corresponding author: Jiang Bian, PhD, College of Medicine, University of Florida, 2197 Mowry Road Suite 122, PO Box 100177, Gainesville, FL 32610-0177.

## Abstract

In this study, we explored the feasibility of using real-world data (RWD) from a large clinical research network to simulate real-world clinical trials of Alzheimer’s disease (AD). The target trial (i.e., NCT00478205) is a Phase III double-blind, parallel-group trial that compared the 23 mg donepezil sustained release with the 10 mg donepezil immediate release formulation in patients with moderate to severe AD. We followed the target trial’s study protocol to identify the study population, treatment regimen assignments, and outcome assessments, and to set up a number of different simulation scenarios and parameters. We considered two main scenarios: (1) a one-arm simulation: simulating a standard-of-care arm that can serve as an external control arm; and (2) a two-arm simulation: simulating both intervention and control arms with proper patient matching algorithms for comparative effectiveness analysis. In the two-arm simulation scenario, we used propensity score matching controlling for baseline characteristics to simulate the randomization process. In the two-arm simulation, higher SAE rates were observed in the simulated trials than the rates reported in original trial, and a higher SAE rate was observed in the 23mg arm than the 10 mg standard-of-care arm. In the one-arm simulation scenario, similar estimates of SAE rates were observed when proportional sampling was used to control demographic variables. In conclusion, trial simulation using RWD is feasible in this example of AD trial in terms of safety evaluation. Trial simulation using RWD could be a valuable tool for post-market comparative effectiveness studies and for informing future trials’ design. Nevertheless, such approach may be limited, for example, by the availability of RWD that matches the target trials of interest, and further investigations are warranted.

## Introduction

Clinical trials, especially randomized controlled trials (RCTs), are critical in the drug discovery and development process to assess the efficacy and safety of the new treatment.^1^ While the rigorously controlled conditions of clinical trials can reduce bias and improve the internal validity of the study results, they also come with the drawbacks of high financial costs and long execution time.^2^ For example, the total cost of developing an Alzheimer’s disease (AD) drug was estimated at $5.6 billion with a timeline of 13 years from the preclinical studies to approval by the Food and Drug Administration (FDA).^3^ Nevertheless, no effective drugs yet have been developed for either treatment or prevention of AD thus far. Strategies that can accelerate the drug development process and reduce costs will not only be of interest to pharmaceutical companies but also ultimately benefit the patients.

Clinical trial simulation (CTS) is a valuable to assess the feasibility, investigate assumptions, and optimize study design before conducting the actual trials.^4,5^ For example, Romero *et al*. conducted a CTS study to explore several design scenarios comparing the effects of donepezil with placebo.^6^ Traditionally, CTS studies use virtual cohorts generated based on pharmacokinetics / pharmacodynamics models of the therapeutic agents, so these cohorts do not necessarily reflect the patients who will use the drugs in the real world. More recently, the trial emulation (i.e., “the target trial”) framework—emulating hypothetical trials to establish the estimation of the casual effects, has attracted significant attention.^7^ For example, Danaei *et al*. emulated a hypothetical RCT used electronic health record (EHR) data from United Kingdom (UK) to estimate the effect of statins for primary prevention of coronary heart disease.^8^ Like many other emulation studies,^7,9,10^ this is essentially a retrospective cohort study, where the authors followed a RCT design to identify unbiased initiation of exposures and eventually to reach an unbiased estimation of the casual relationship. Combining the ideas from CTS and trial emulation, a simulation study using real-world data (RWD) to test different assumptions (e.g., different drop-out rates) and trial designs (e.g., different eligibility criteria) could provide insights on the effectiveness and safety of the treatments to be developed in a real-world setting that reflect the patient populations who will actually use the treatment.

In this study, we explored the feasibility of using RWD from the OneFlorida Clinical Research Consortium—a clinical data research network funded by the Patient-Centered Outcomes Research Institute (PCORI) contributing to the national Patient-Centered Clinical Research Network (PCORnet)—to simulate a real-world AD RCT as a use case. We considered two main scenarios: (1) a one-arm simulation: simulating a standard-of-care arm that can serve as an external control arm; and (2) a two-arm simulation: simulating both intervention and control arms with proper patient matching algorithms for comparative effectiveness analysis.

## Results

### Computability of eligibility criteria in the original trial (i.e., NCT00478205)

In total, there are 36 eligibility criteria in trial NCT00478205, where 17 are inclusion and 19 are exclusion criteria. However, not all criteria are computable against the OneFlorida patient database: (1) 11 are not computable, and (2) 7 are partially computable (i.e., a part of the criterion is not computable). Similar to what we have found in our prior study^11^, the common reasons for not computable criteria are (1) data elements needed for the criterion do not exist in the source database (e.g., “*A cranial image is required, with no evidence of focal brain disease that would account for dementia*.”), or (2) the criterion asked for subjective information either from the patient (e.g., *“Patients who are unwilling or unable to fulfill the requirements of the study*.*”) or the investigator (e*.*g*., *““Clinical laboratory values must be within normal limits or, if abnormal, must be judged not clinically significant by the investigator*.”). When a criterion is not computable, we consider all candidate patients met that criterion (e.g., they are all willing and able to “*fulfill the requirements of the study*”).

### Characteristics of the target, study, and trial not eligible populations from OneFlorida

Overall, a total of 90 and 2,048 patients were identified as the effective target populations in OneFlorida for the 23 mg arm and 10 mg arm, respectively. Among them, 38 and 782 met the eligibility criteria of the original target RCT for the two arms, respectively. ***Table 2*** shows the demographic characteristics and SAE statistics of the original trial population as well as the effective target population (TP), study population (SP), and trial not eligible population (NEP) from OneFlorida.

**Table 1.**
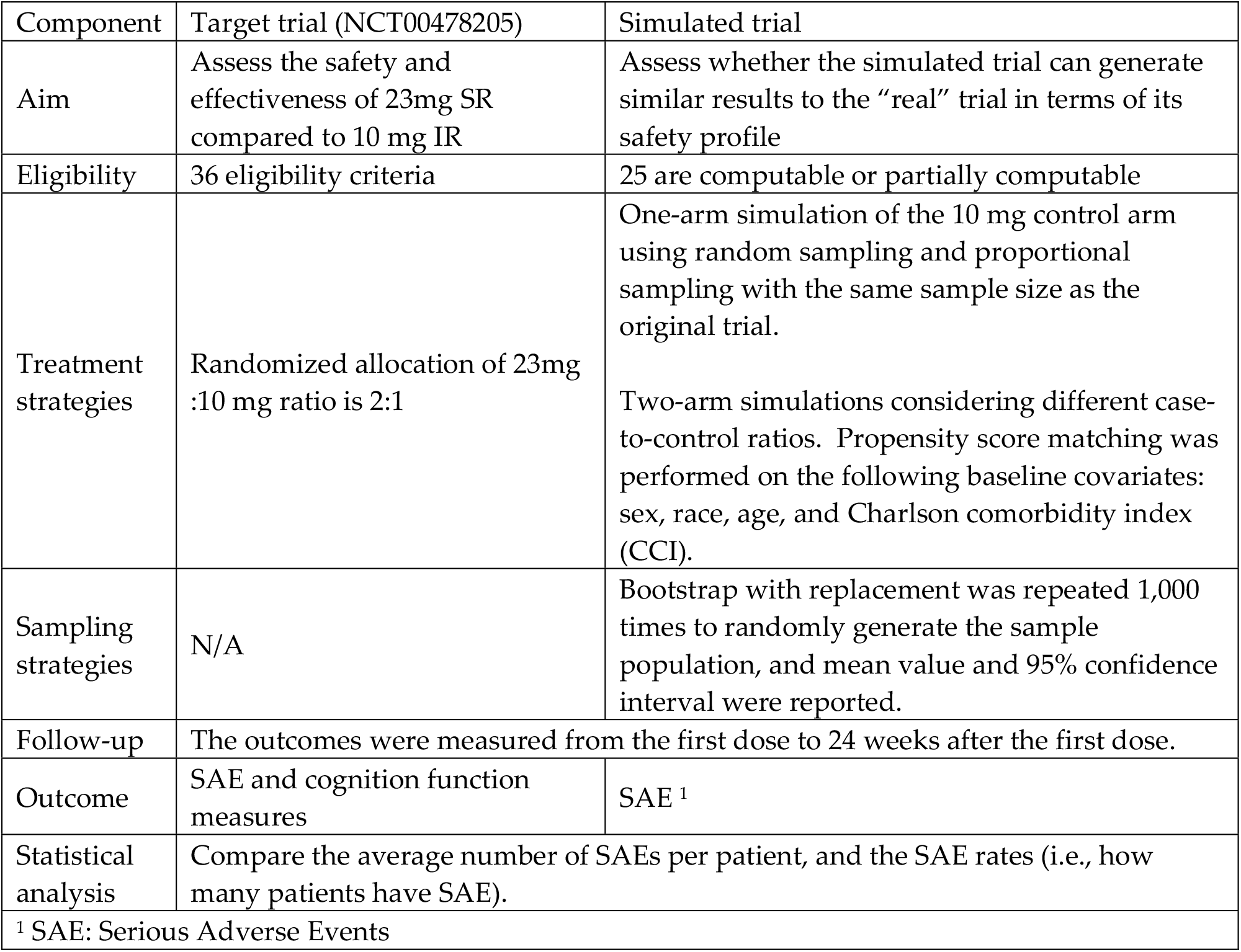
Overall study design of the simulated trial in comparison with the original trial.

**Table 2.** Population characteristics and SAE statistics of the target trial vs. TP, SP, and NEP from OneFlorida.

|  | 23mg Arm |  |  |  | 10mg Arm |  |  |  |
| --- | --- | --- | --- | --- | --- | --- | --- | --- |
|  | Original Trial<sup>1</sup> | Overall TP<sup>2</sup> | Overall SP<sup>3</sup> | Overall NEP<sup>4</sup> | Original Trial | Overall TP<sup>5</sup> | Overall SP<sup>6</sup> | Overall NEP<sup>7</sup> |
| # of Subject | 963 | 90 | 38 | 52 | 471 | 2,048 | 782 | 1,266 |
| Age Mean (SD) | 73.9<br>(8.53) | 74.3<br>(9.01) | 73.3<br>(9.01) | 81.6<br>(12.37) | 73.8<br>(8.56) | 73.4<br>(11.0) | 74.2<br>(9.67) | 77.1<br>(11.8) |
| Gender |  |  |  |  |  |  |  |  |
| Male | 356<br>(37.0%) | 24<br>(26.7%) | 10<br>(26.3%) | 14<br>(26.9%) | 177<br>(37.6%) | 727<br>(35.5%) | 234<br>(29.9%) | 493<br>(38.9%) |
| Female | 607<br>(63.0%) | 66<br>(73.3%) | 28<br>(73.7%) | 38<br>(73.1%) | 294<br>(62.4%) | 1321<br>(64.5%) | 548<br>(70.1%) | 773<br>(61.1%) |
| Race<sup>e</sup> |  |  |  |  |  |  |  |  |
| White | 708<br>(73.5%) | 63<br>(70.0%) | 25<br>(65.8%) | 38<br>(73.1%) | 346<br>(73.5%) | 829<br>(40.5%) | 280<br>(35.8%) | 549<br>(43.4%) |
| Asian/Pacific | 161<br>(16.7%) | 0<br>(0%) | 0<br>(0%) | 0<br>(0%) | 87<br>(18.5%) | 22<br>(1.1%) | 11<br>(1.4%) | 11<br>(0.9%) |
| Hispanic | 67<br>(7.0%) | 15<br>(16.7%) | 4<br>(10.5%) | 11<br>(21.2%) | 26<br>(5.5%) | 440<br>(21.5%) | 192<br>(24.6%) | 248<br>(19.6%) |
| Black | 22<br>(2.3%) | 11<br>(12.2%) | 4<br>(10.5%) | 7<br>(13.5%) | 9<br>(1.9%) | 380<br>(18.6%) | 126<br>(16.1%) | 254<br>(20.1%) |
| Other | 5<br>(0.5%) | 1<br>(1.1%) | 5<br>(13.2%) | 7<br>(13.5%) | 3<br>(0.6%) | 377<br>(18.4%) | 173<br>(22.1%) | 204<br>(16.1%) |
| CCI<sup>f</sup> | N/A | 1.54 | 1.32 | 1.53 | N/A | 2.36 | 1.64 | 2.36 |
| Mean SAE<sup>g</sup> | 0.15 | 1.89 | 0.92 | 2.60 | 0.14 | 1.68 | 0.64 | 2.59 |
| # of patients with<br>≥ 1 SAE | 45 (9.6%) | 20<br>(22.2%) | 4 (10.5%) | 16<br>(30.7%) | 80 (8.3%) | 573<br>(28.0%) | 121<br>(15.5%) | 452<br>(35.7%) |

<sup>1</sup>Reported in the original trial on ClinicalTrials.gov
<sup>2</sup>TP: Target population—patients who (1) had the disease of interest (i.e., AD), and (2) had used the study drug (i.e. donepezil) for a specific time period according to the study protocol.
<sup>3</sup>SP: Study population—patients in the TP who met the computable eligibility criteria of the original trial.
<sup>4</sup>NEP: Trial not eligible population—patients in the TP who did NOT meet the eligibility criteria of the original trial.
<sup>5</sup>The original trial reported Hispanic as a race, thus, we followed the same convention to make sure the results are comparable even though race and ethnicity are two different fields in OneFlorida.
<sup>6</sup>Charlson Comorbidity Index.
<sup>7</sup>Mean SAE: average number of SAEs per patient.

For demographic characteristics, relative to the target RCT population, we observed a large difference in race in our OneFlorida population (all p-values of race group comparisons were smaller than 0.05). OneFlorida had more Hispanics (10.5% - 24.6% vs. 5.5% - 7%) and Blacks (10.5% - 20.1% vs. 1.9% - 2.3%), but less Whites (35.8% - 73.1% vs. 73.5% - 73.5%) or Asian/Pacific islanders (0% - 1.4% vs. 16.7% - 18.5%). The age distributions were similar across all populations. For clinical variables, we calculated the Charlson Comorbidity Index (CCI) of the various populations from OneFlorida. Smaller CCIs were observed in the SP compared with the TP for both arms (p<0.05), and a smaller CCI was observed in the 23mg arm compared with the 10mg arm (p<0.05). Our primary outcomes of interest in this analysis were SAEs. Thus, we calculated the mean SAE (i.e., the average number of SAEs per patient) and the number of patients who had more than 1 SAE during the study period. For both 23mg and 10mg arms, the mean SAE and the number of patients with SAEs were the largest in the TP, followed by the SP, and then the original trial. Consistent with the original trial, populations derived from the OneFlorida data in the 23mg arm have higher numbers of mean SAE and more patients with SAE compared with the 10mg arm.

### Standard-of-care control arm (i.e., one-arm) simulation

We first simulated the control arm of the original trial (i.e., the 10mg stand-of-care arm). ***Table 3*** displays the demographics and SAE outcomes in the simulated control arms. Here, we reported the mean value and 95% confidence interval of all 1,000 bootstrap samples. Two different sampling approaches were used: (1) random sampling, and (2) proportional sampling accounting for race distribution. When using the random sampling approach, compared with the control arm in the original trial, higher mean SAE and SAE rates were observed, in addition to discrepancies in demographic variables. When using proportional sampling, the results were closer and more consistent with the original trial. Notably, the SAE rates in the simulated control were similar to the SAE rates from the original control (8.9% vs. 8.3%), and a z-score test for population proportion had a p-value of 0.75, suggesting there were no significant differences between the two SAE rates. In addition to SAE rates and mean SAE, we also explored the SAE event rates in the simulated control arms stratified by the SAE category reported in the original trial. Compared with the control arm in the original trial, the simulated control arms have larger SAE rates in most categories.

**Table 3.** One-arm simulation results for the 10mg control arm.

|  | Original control arm | Simulated control |  |
| --- | --- | --- | --- |
|  |  | Random sampling | Proportional sampling |
| Number of subjects | 471 | 400 | 240 |
| Mean age | 73.8 | 78.5±0.1 | 78.2±0.1 |
| Gender |  |  |  |
| Female | 62.4% | 70.1%±0.1% | 66.1%±0.1% |
| Male | 37.6% | 29.9%±0.1% | 33.9%±0.1% |
| Race |  |  |  |
| White | 73.5% | 28.1%±0.1% | 73.3%±0.1% |
| Black | 1.9% | 16.0%±0.1% | 2.1%±0.1% |
| Hispanic | 5.5% | 24.5%±0.1% | 5.4%±0.1% |
| Asian & Other | 19.1% | 31.3%±0.2% | 19.2%±0.1% |
| SAE<sup>1</sup> rates (patient with ≥ 1 SAE) | 8.3% | 15.5%±0.1% | 8.9%±0.1% |
| Mean SAE (average SAE per patient) | 0.14 | 0.64±0.01 | 0.48±0.01 |
| SAE event rates by category |  |  |  |
| Blood and lymphatic system disorders | 0/471 (0.00%) | 4.9%±0.1% | 3.2%±0.1% |
| Cardiac disorders | 6/471 (1.23%) | 2.9%±0.1% | 2.0%±0.1% |
| Eye disorders | 1/471 (0.2%) | 0 | 0 |
| Gastrointestinal disorders | 2/471 (0.4%) | 7.0%±0.1% | 5.9%±0.1% |
| General disorders | 3/471 (0.6%) | 1.7%±0.1% | 1.1%±0.1% |
| Hepatobiliary disorders | 2/471 (0.4%) | 1.2%±0.1% | 1.9%±0.1% |
| Infections and infestations | 9/471 (1.9%) | 5.8%±0.1% | 4.7%±0.1% |
| Injury, poisoning and procedural complications | 8/471 (1.7%) | 3.2%±0.1% | 3.0%±0.1% |
| Musculoskeletal and connective tissue disorders | 2/471 (0.4%) | 0.7%±0.1% | 0.8%±0.1% |
| Neoplasms benign, malignant and unspecified | 2/471 (0.4%) | 0 | 0 |
| Nervous system disorders | 15/471 (3.3%) | 6.6%±0.1% | 4.8%±0.2% |
| Psychiatric disorders | 11/471 (2.3%) | 3.3%±0.1% | 2.5%±0.1% |
| Renal and urinary disorders | 2/471 (0.4%) | 4.2%±0.1% | 1.9%±0.1% |
| Reproductive system and breast disorders | 0 | 0 | 0 |
| Respiratory, thoracic and mediastinal disorders | 1/471 (0.2%) | 1.0%±0.1% | 1.4%±0.1% |
| Vascular disorders | 1/471 (0.2%) | 0.7%±0.1% | 0.3%±0.1% |
| <sup>1</sup>SAE: severe adverse events |  |  |  |

### Two-arm trial simulation

Because the proportional sampling had better performance in the one-arm simulation, we used this sampling strategy in the two-arm simulation to match the race distribution, and tested two scenarios of different matching ratios (i.e., proportional 1:1 matching and proportional 1:3 matching). However, since there is no Asian/Pacific in our study population who used 23mg donepezil, all Asians in the original trial and the simulated control arm were grouped into “other”, and the sample size of the 23mg arm was set at 30 (because of the limited number of 23mg patients in the OneFlorida data). Table 4 shows our two-arm simulation results, where we show the average and 95% CI of all variables for the simulation arms across all 1,000 bootstrap samples. In both matching scenarios, the mean SAE and SAE rates were higher in the 23mg arm than in the 10mg arm, which is consistent with the original trial. However, the variance for both SAE outcomes for the 10mg arm are higher in the 1:1 matching scenario than in the 1:3 matching scenario, as the sample size for the 10mg arm in the 1:3 matching scenario is much bigger. Because of the sample size difference, estimates from the 1:3 matching scenario should be more reliable. Consistent with the original trial, patients in the 23mg arm have higher event rates in most of the SAE categories compared to the patients in the 10mg arm. Note that we observed no SAE events in several categories in our simulation, especially in the 23mg arm, due to the limited sample size.

**Table 4.**
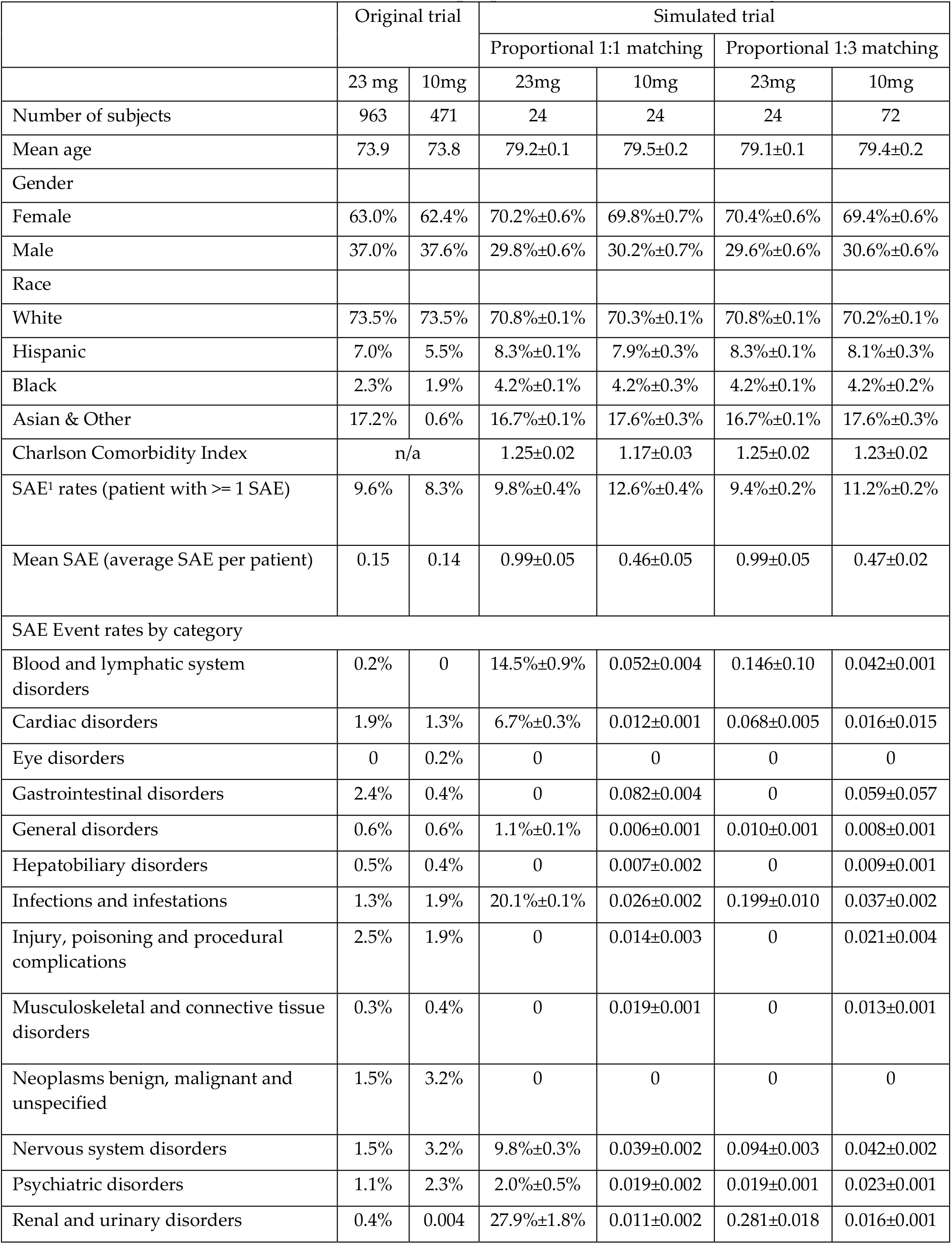
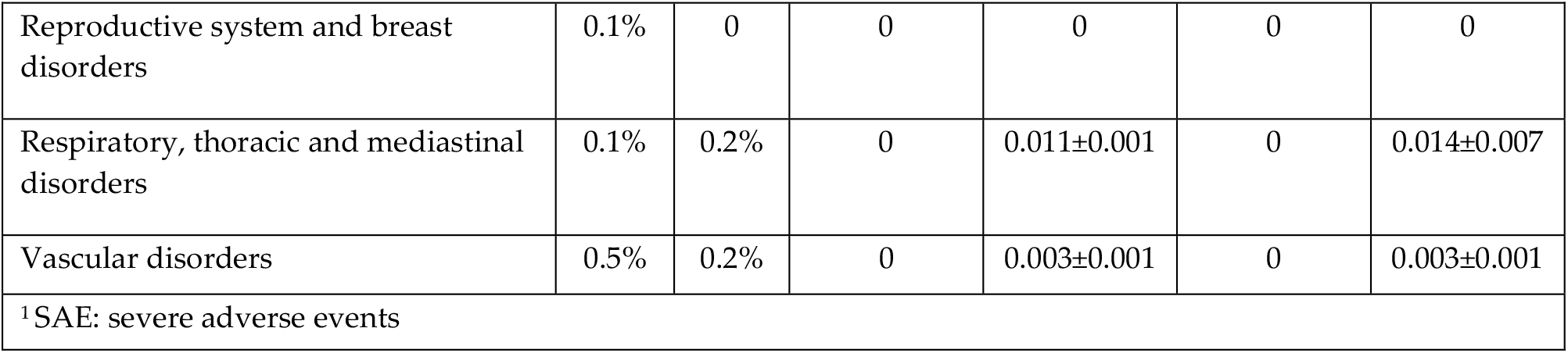
Results of the two-arm simulation for proportional 1:1 and 1:3 matching.

Finally, we conducted an additional experiment to simulate patients who withdrew from the trial. In the original trial, among the 963 and 471 patients from each arm, 296 (30%) and 87 (18%) patients discontinued the study for various of reason, respectively. Among the dropouts, 182 and 39 patients discontinued due to AEs. We simulated the dropouts by (1) randomly removing 18% in the 10mg group and 30% in the 23mg in our simulations; and (2) removing the patient after his or her first AE using the same proportion as the original trial (i.e., due to small sample size, we did not simulate this scenario for the two-arm simulation). The results are displayed in **Supplementary Table 3** and **Supplementary Table 4**. For the random dropout scenario, similar SAE rates and smaller mean SAE were observed across all scenarios. For example, in the control arm of the random dropout scenario, mean SAE decreased from 0.64 to 0.19 and 0.23 in the two different dropout simulations, while in the two-arm simulation, the mean SAE for the 10mg arm were 0.22 and 0.19 in the two scenarios, both were much lower than simulations without dropout (0.46 and 0.47 respectively). However, the effects of dropout were mostly observed in control arms, the SAE rates and mean SAE remained the same for the 23mg arm before and after dropout simulation. In the AE-based dropout scenario, both smaller SAE rates and smaller mean SAE were observed: the SAE rates decreased from 8.8% to 7.3% and the mean SAE decreased from 0.64 to 0.23, where the mean SAE estimate was closer to the original trial results.

## DISCUSSION

In this work, we simulated an AD RCT utilizing RWD from the OneFlorida network—a large clinical data research network, considering three different simulation scenarios. In the one-arm simulation scenario, we attempted to simulate an external control arm for the original trial. We demonstrated that we could achieve similar estimate of SAE rates as the original trial when proportional sampling accounting for race distribution was used; and the statistics of the simulated control arm were stable across all bootstrap simulation runs, which suggests that using RWD we can robustly simulate the “*standard of care*” control arm. In the two-arm comparative effectiveness simulations, we used propensity score matching on baseline characteristics to simulate the randomization process. It has been demonstrated that propensity score matching could reduce bias in the estimate of the treatment response,^12–15^ and in our study, we successfully simulated two groups of patients that have similar age, sex, race, and CCI distributions using propensity score matching. However, the SAE outcomes in the simulated trial were still different from the original trial for various reasons (1) the original trial was conducted in research settings, while RWD data reflect patients the real-world clinical settings. The total time at risk for SAEs in our simulated cohort may be longer than the original trial, because in clinical trials, patients may withdraw from the study once experiencing an SAE, while patients in real-world setting may not. This is demonstrated by our simulation of dropouts, which achieved smaller number of SAEs and closer results to the original trial; (2) sample size issue, where 23mg donepezil has not been the standard-of-care for AD in the real-world, leading to considerably fewer patients in the 23mg arm. For the two-arm simulation, we conducted a post-hoc power analysis with the SAE rates and mean SAE. Assuming a significant level at 0.05, a 65% power were achieved; and (3) although propensity score matching derived two simulation arms that are comparable, we were unable to compare it directly to the study population in the original trials as the data for calculating propensity scores were not available from the original trial. For example, the switching to the 23mg treatment after receiving at least 3 months of the 10mg does not occur at random in a real-world setting, but based on clinical guidelines; and indeed, we found that patients in the 23mg arm have a longer history of diagnosis (i.e., mean of days between first diagnosis and first prescription in 23mg arm is 398 days vs. 128 days in the 10mg arm). Therefore, in our two-arm simulation, there may be residue selection bias causing a difference between the two populations; Nevertheless, this is an issue of using observational data in general; even though we can simulate randomization, e.g., through propensity score matching; but trial simulations cannot replace RCTs. In addition, there are still gaps, especially data gaps in RWD, that also contributed to the differences between our simulation results and the original trial results. Future studies are warranted to identify strategies to fill these gaps.

While simulating the original AD trial followed the study protocol in **Table 1**, we found it is difficult to replicate all the eligibility criteria of the original trial. Out of the 36 eligibility criteria, only 25 of them were computable or partially computable against the OneFlorida data. Since these criteria were used to weed out patients who are unlikely to complete the protocol (e.g., due to safety concerns), ignoring some of the criteria (not computable eligibility criteria) could potentially explain some of the increases either in the mean SAE or the SAE rates. One strategy for future simulation studies is to classify each of the eligibility criteria based on their clinical importance to the simulation study and the endpoints (i.e., effectiveness or safety) related with the criterion. By doing so, we can adjust the eligibility criteria and customize the simulation based on questions of interest. For example, efficacy-related criteria may have very small impact on a trial that is focused on examining safety and toxicity; so simulations of such trials can loosen the restrictions on efficacy-related criteria. Nevertheless, as all the patients we identified in the OneFlorida data have taken the study drugs of interest (i.e., different dosages of donepezil), they should all have been eligible to the original trial in an ideal world, where the trial participants truly reflect target population (i.e., higher trial generalizability).

Our findings are consistent with previous literature on clinical trial generalizability.^16–19^ More SAEs were observed in real-world settings. In our data, the overall number of patients who had SAEs and the average number of SAEs per patient were (1) the highest in the effective target population (i.e., patients who took donepezil for AD), which is the population who actually used the medication in real-world settings, but also (2) higher in the study population—patients who used donepezil for AD and also met the original trial’s eligibility criteria. Some of the differences may be due to the incomputable eligibility criteria (e.g., general physical health deterioration) that we cannot account for, but it is also possible that the original trial samples did not adequately reflect the TP and thus there might be treatment effect heterogeneity across patient subgroups, not captured by the original trial. In the two-arm simulations, large variances were observed, especially when the matched sample size was small. This may also indicate the heterogeneous treatment effects of donepezil when applied to different patient subgroups in real-world settings.

Our study demonstrated the feasibility of trial simulation using RWD, especially when simulating external standard-of-care control arms. Our one-arm simulation provided stable and robust estimates and sufficient sample sizes to compare with the original trial’s control arm. The SAE rates observed in the simulated control arm with proportional sampling were very close to what was reported in the original trial. The mean SAE per patient and SAE event rates, however, were larger in the simulated control arms, which suggested that, in a real-world setting, the patients who experienced SAEs tend to have more occurrences of SAEs. On the other hand, the two-arm simulation, although it provided insights, was not entirely successful. Although the randomization process was effectively simulated by using propensity score matching, the outcome measures were very different from the original trial. The reasons for the differences could be multi-fold (e.g., research setting vs. real-world clinical setting, difference in sample size, overly restrictive eligibility criteria that limits the generalizability of the original trial), but cannot be explored due to limited data reported by the original trial (i.e., no patient-level data is available). When we simulated dropouts using information from the original trial, we observed similar SAE rates and mean SAE compared to the original trial in both random dropout or AE-based dropout strategies, especially in the control arms. This suggests that the additional simulation scenarios have led to results more comparable to the cohorts in the original trial. However, due to the small sample size in the 23mg arm, we were only able to simulate random dropout for the two-arm simulation, and the SAE measurements did not change much comparing to the simulations without dropout. Future studies with sufficient sample sizes could conduct more sophisticated analysis based on treatment delay and adherence.

Compared with trial emulation, which focus on making the target trial explicitly characterized with a defined protocol, our approach takes the advantage of having observational RWD, where different trial protocols (i.e., simulation scenarios) with different study designs can be readily tested. For example, in our current study, there are several potential simulation points that can be further tuned. First, the sample size of each arm can be adjusted. In the one arm simulation (i.e., the control arm of 10mg donepezil), we choose the same sample size as the original trial, but it can be adjusted to increase power. Second, the eligibility criteria of selecting study population could also be adjusted to test different hypotheses. For example, we can adjust the eligibility criteria in the trial simulation process to assess how the original trial results may be generalized into real-world target population, and provide insights on how to balance internal validity while retaining good external validity.^17,20^ Third, different scenarios of dropout may be simulated. The dropout rate and timing can be varied, so that it can be used to simulate different patient population. Further, we can also explore whether other dropout reasons such as lack of recovery and lack of access to care can be simulated based on real-world data. Many other potential simulation scenarios can be tested such as varying the 3-month lead time for switching from 10mg to 23mg. In this current work, as our main goal was to establish the feasibility of such simulation approach, we only conducted limited number of major simulation scenarios (e.g., we used two different sampling scenarios using different intervention arm vs. control arm ratios). In future work, informed by literature, we shall systematically simulate the different trial design scenarios, which can (1) provide critical information on the comparative effectiveness of the interventions in real-world settings, but also (2) better inform the study designs of future clinical trials. Last but not least, the one-arm simulation is as important as the two-arm simulation, even though it does not provide comparative effectiveness results of the intervention. In addition to informing future design of control arms, one-arm simulation allows us to utilize readily available RWD of patients taking the standard-of-care (SOC) to determine SOC’s treatment effectiveness and safety profile and consider different study protocols and scenarios. The demonstrated feasibility of one-arm simulation is a building block towards the potential of using RWD to generate synthetic and external controls for clinical trials, leading to significant cost savings.^21^ Nevertheless, other issues with RWD such as its data quality (e.g. missing key measures of endpoints) and the inherent biases exist in observational data, warrant further investigations.

There are some other limitations in this study. First, we only looked at one original trial for one medication (i.e., donepezil). Simulations on different drugs and diseases may have different results. Second, the population who took the 23mg form in our data is very small (even though the overall OneFlorida population is large with more than 15 million patients), where we only identified 38 patients who took the 23mg donepezil and met the eligibility criteria of the original trial. The 23mg donepezil form was approved by the FDA in 2012, so it is still a relatively new drug on the market, and following its approval, the clinical utility of the 23mg form was called into question because of its limited effectiveness and higher rates of adverse events.^22^ The current practice of using the donepezil 23mg form is reserved for AD patients who have been on stable donepezil 10 mg form for at least 3–6 months with no significant improvement,^31,32^ which limited its use in real-world clinical practice. In addition, patients who switched to the 23mg treatment may have different characteristics in characteristics that we did not account for in this analysis. Third, we found some of the SAEs (e.g., abnormal behavior, presyncope) reported in the trial’s results cannot be mapped to any AE terms in CTCAE, and the definitions of AEs in the original trial were unavailable, which increased the difficulty of accurately accounting for all SAEs. Further, even though trials’ SAEs reported in ClinicalTrials.gov largely follow the Medical Dictionary for Regulatory Activities Terminology (MedDRA), not all reported SAEs were correctly defined in the trial results. For example, we found “*Back pain*” and “*Fall*” were defined as SAEs in the original AD trial we modeled. However, in CTCAE, there is no corresponding category 4 or 5 definition for them. More effort is needed to consistently model SAEs reported in clinical trials. Finally, because of data limitations, we were not able to assess the effectiveness of AD treatment (e.g., AD end points such as Mini-Mental State Examination and Severe Impairment Battery are not readily available in structure EHR data, but may exist in clinical narratives). Thus, we only examined safety outcomes in our current study. This may also contribute to the different results we obtained from our simulation compared with results from the original trial as the original trial was designed and powered with primary efficacy-based outcomes. Future studies that explore the use of advanced natural language processing (NLP) methods to extract these endpoint measurements from clinical notes will be important. Further, variables extracted from clinical notes with NLP could also be used to render some of the incomputable eligibility criteria computable.

In conclusion, in this study, we investigated the feasibility of using the existing patient records to simulate clinical trials using an Alzheimer’s disease trial (i.e. NCT00478205) as the use case. We examined two main simulation scenarios: (1) a one-arm simulation: simulating the standard-of-care arm that can serve as an external control arm; and (2) a two-arm simulation: simulating both intervention and control arms with proper patient matching algorithms for comparative effectiveness analysis. We have also considered a number of different simulation parameters such sampling strategies, matching approaches, and dropout scenarios. In the case study, our simulation can robustly simulate “standard of care” control arms (i.e. the 10mg donepezil arm) in terms of safety evaluation. However, trial simulation using RWD may be limited by the availability of RWD that matches the target trials of interest and may not yield reliable and consistent results if the sample sizes of the interventions of interest (i.e. we found few patients were prescribed the 23mg donepezil) are limited from the real-world databases. Further investigations on this topic are warranted, especially how to address the data quality issues (e.g. using NLP to extract more complete patient information) and reduce inherent biases (e.g. more advanced matching methods to tackle the problems of high dimensionality, nonlinear/nonparallel treatment assignment, and other complex confounding situations^25^) in observational RWD. Last but not least, it will also be beneficial to have access more complete information (e.g. de-identified individual-level trail participant data) of the target trials, so that more realistic simulation settings can be explored.

## Methods

### The target Alzheimer’s disease (AD) trial and its characteristics

Although there is no cure for AD yet, the U.S. FDA approved two classes of medications: (1) cholinesterase inhibitors, and (2) memantine, to treat the symptoms of dementia. Donepezil (Aricept®), a cholinesterase inhibitor, was the most widely tested AD drug and approved for all stages of AD. The target trial NCT00478205^26^ is a Phase III double-blind, double-dummy, parallel-group comparison of 23 mg donepezil sustained release (SR) with the 10 mg donepezil immediate release (IR) formulation (marketed as the standard-of-care) in patients with moderate to severe Alzheimer’s disease. Patients who have been taking 10 mg IR (or a bioequivalent generic) for at least 3 months prior to screening were recruited. The original trial consisted of 24 weeks of daily administration of study medication, with clinic visits at screening, baseline, 3 weeks (safety only), 6 weeks, 12 weeks, 18 weeks, and 24 weeks or early termination. Patients received either 10 mg donepezil IR in combination with the placebo corresponding to 23 mg donepezil SR, or 23 mg donepezil SR in combination with the placebo corresponding to 10 mg donepezil IR. A total of 471 and 963 patients were enrolled from approximately 200 global sites (Asia, Oceania, Europe, India, Israel, North America, South Africa, and South America). The results of the original trial yielded that donepezil 23 mg/d was associated with greater benefits in cognition compared with donepezil 10 mg/d and led to the FDA approval of the new 23 mg dose form for treatment of AD in 2010,^27^ despite the debate on whether the 2.2 point of cognition improvement (on a 100 point scale) over the 10 mg dose form is sufficient.^22,28^

In our simulation, we followed the detailed study procedures outlined in their published article^27^ to formulate our simulation protocol, including the treatment regimen, population eligibility, and follow-up assessments for SAEs. **Table 1** describes how the original trial design was followed in our simulation.

### Real-world patient data (RWD) from the OneFlorida network

The OneFlorida data contain robust longitudinal and linked patient-level RWD of ∼15 million (>60%) Floridians, including data from Medicaid claims, cancer registries, vital statistics, and EHRs from its clinical partners. As one of the PCORI-funded clinical research networks in the national PCORnet, OneFlorida includes 12 healthcare organizations that provide care through 4,100 physicians, 914 clinical practices, and 22 hospitals, covering all 67 Florida counties. The OneFlorida data is a Health Insurance Portability and Accountability Act (HIPAA) limited data set (i.e., dates are not shifted and location data are available) that contains detailed patient characteristics and clinical variables, including demographics, encounters, diagnoses, procedures, vitals, medications, and labs.^29^ We focused on the structured data immediately available to us formatted according to the PCORnet common data model (PCORnet CDM).^30^

### Cohort identification: the target population, the study population, and the trial not eligible population

From the OneFlorida data, we identified three populations: the target population (TP), the study population (SP), and the trial not eligible population (NEP) for the target trial following the process shown in **Figure 1 Panel a**, and the relationship between these populations are displayed in **Figure 1 Panel b**. The *true* target population should be those that will benefit from the drug, thus, should be broader as patients with AD in general. However, as patients who were not treated with donepezil in real-world would not have any safety or effectiveness data of the drug in RWD, the effective target population of interest is a constrained subset—patients who (1) had the disease of interest (i.e., AD), and (2) had used the study drug (i.e., donepezil) for a specific time period according to the study protocol. The 10mg donepezil is only in immediate release (IR) form while the 23mg donepezil is exclusively in sustained release (SR) form, so we used the corresponding RxNorm concept unique identifier (RXCUI) and the National Drug Code (NDC) to identify the two groups (i.e., 10mg vs. 23mg) of patients in our data.^31,32^ We then identified the study population (i.e., patients who met both the TP criteria and the trial eligibility criteria) and trial not eligible population (i.e., patients who meet the TP criteria but do not meet the trial eligibility criteria) by applying the eligibility criteria of the target trial to the TP. To do so, we analyzed the target trial’s eligibility criteria and determined the computability of each criterion. A criterion is computable when its required data elements are available and clearly defined in the target patient database (i.e., the OneFlorida data in our study). Then, we manually translated the computable criteria into database queries against the OneFlorida database. We assumed that all patients met the non-computable criteria (e.g., “*written informed consent*”), which is a limitation of our study. The full list of eligibility criteria and their computability are listed in the **Supplementary Table 2**. We first decomposed each criterion (e.g., “*Patients with dementia complicated by other organic disease or Alzheimer’s disease with delirium*”) into smaller study traits (e.g., “dementia complicated by other organic disease” and “Alzheimer’s disease with delirium”). We then checked whether each of the study trait is computable based on the OneFlorida data as shown in **Supplementary Table 2**. We then used the computable study traits to determine patients’ eligibility. Many of the incomputable study traits are not clinically relevant for our studies (e.g., “*No caregiver available to meet the inclusion criteria for caregivers*.”). Nevertheless, how computability of these study traits affect the trial simulation results – a limitation of our current study – warrant further investigations in future studies.

**Figure 1.**
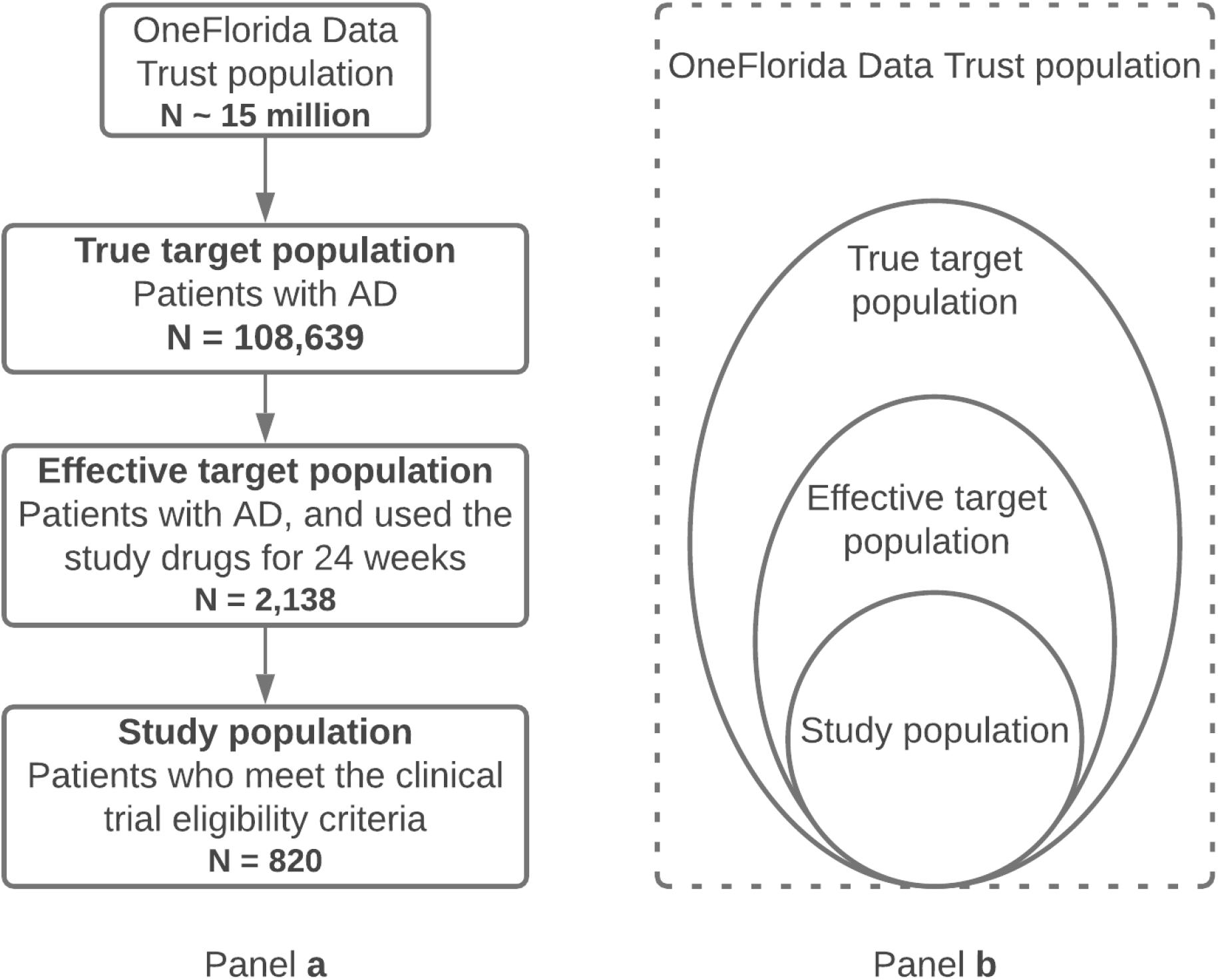
The cohort identification process for the target, study, and trial not eligible populations.

### Definition and identification of serious adverse events (SAE) from EHRs

The target trial used Severe Impairment Battery (SIB) and the Clinician’s Interview-Based Impression of Change Plus Caregiver Input scale (CIBIC+; global function rating) to assess the efficacy of donepezil in AD patients. Because these effectiveness data are not readily available in the structured EHR data, we focused on assessing drug safety in terms of the occurrences of SAEs. To define an SAE, we followed the FDA^33^ definition of SAEs and the Common Terminology Criteria for Adverse Events (CTCAE) version 5 – a descriptive terminology for Adverse Event (AE) reporting. In CTCAE, an AE is any “*unfavorable and unintended sign, symptom, or disease temporally associated with the use of a medical treatment or procedure that may or may not be considered related to the medical treatment or procedure*,” and the AEs are organized based on the System Organ Class (SOC) defined in Medical Dictionary for Regulatory Activities (MedDRA^34^). CTCAE also provides a grading scale for each AE into Grade 1 (mild), Grade 2 (moderate), Grade 3 (severe or medically significant but not immediately life-threatening), Grade 4 (life-threatening consequences), and Grade 5 (death).

We mapped each reported SAE in the trial results section of the target trial NCT00478205 on ClinicalTrails.gov at https://www.clinicaltrials.gov/ct2/show/results/NCT00478205 to the CTCAE term and identified the severity based on the CTCAE grading scale. We considered an AE as SAE if it meets the criteria for Grade 3/4 (results in hospitalization), and Grade 5 (death). As shown in **Figure 2**, to count as a SAE related to donepezil, the SAE event has to occur within 24 weeks after the first donepezil prescription (which is the same follow-up period as the original trial). Note that we excluded chronic conditions that happened before the study, for example, different types of cancer.

**Figure 2.**
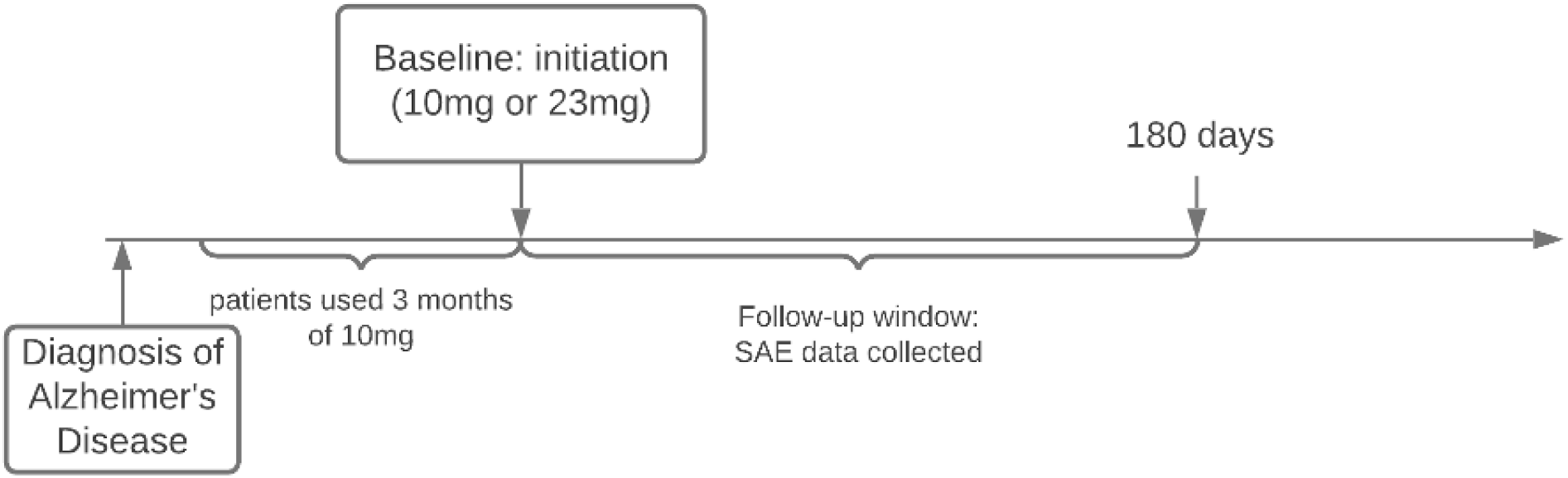
Follow-up window for serious adverse events (SAEs) related to treating Alzheimer’s disease with donepezil.

### Trial simulation

***Table 1*** shows our design of the simulated trial corresponding to the original target trial. Based on the calculation from the original trial^27^, a sample size of 400 and 800 were needed for the 10mg and 23mg arms, respectively. We first simulated the control arm of the standard therapy (i.e., the 10 mg arm of the original trial), where we have a sufficiently large sample size from the OneFlorida data. We designed our simulation based on the sample size of the arm in the original trial (N=400), and tested two different sampling approaches: (1) random sampling, and (2) proportional sampling controlling for race distribution.

Even though we did not find a sufficient number of patients who took 23mg donepezil in our data, we still simulated both case-control arms using the same sampling strategy in the one-arm simulation that yielded the closest effect sizes compared with the original trial. We explored two different scenarios with different sample sizes: (1) the ratio of the number of subjects in the 23mg arm to the 10mg arm was set as 1 to 1, and (2) the ratio was set as 1 to 3. Because of the limited number of individuals who took the 23mg form, we can only increase the number of subjects in the 10mg arm in the second sample size scenario. We used propensity score matching (PSM) to simulate randomization. The variables used for PSM included age, gender, race, and Charlson comorbidity index (CCI, i.e., as a proxy for baseline overall health of the patient) prior to baseline. Specifically, we fitted a logistic regression model using different treatment (i.e., case vs. control) as the outcome variable and age, gender, race, and CCI as covariates to generate the logistic probabilities of propensity scores individuals in the two comparison groups and then used the nearest neighbor method to carry out the mapping process. The two arms were matched with the propensity scores with a 1:1 or 1:3 ratio.

Specifically, we first used proportional sampling to extract a sample of patients for the 23mg study population using the same race distribution as in the original trial, and then identified a matched sample for the 10mg study population using PSM. We then calculated the SAEs in the 10mg vs. 23mg arms as the safety outcomes. The simulation process was performed 1,000 times with bootstrap sampling with replacement, and the mean value and 95% confidence interval (CI) of each bootstrap sample were calculated to generate the overall estimates. We focused on comparing the average number of SAE per patient, the overall SAE rates (i.e., how many patients had SAEs), and stratified the analysis by major SAE categories according to the CTCAE guideline. The effects of PSM were evaluated by examining the distributions of propensity scores using jitter plot (**Supplementary Table 1** and **Supplementary Figure 1**).

## Data Availability

The data used in this work are available from the OneFlorida Clinical Research Consortium upon request and committee review.

## Data availability statement

OneFlorida data can be requested at https://onefloridaconsortium.org/front-door/; Since OneFlorida data is a HIPAA limited data set, a data use agreement needs to be established with the OneFlorida network.

## Code availability statement

The data processing and analysis were conducted using R version 4.0.2. The code used in this work is available upon reasonable request for academic purposes.

## Acknowledgements

This work was supported in part by NIH grants R21AG068717, R21AG061431, and UL1TR001427. The content is solely the responsibility of the authors and does not necessarily represent the official views of the NIH.

## Author contributions

ZC, HZ, and JB designed the initial concepts; ZC and HZ carried out the analysis; ZC, HZ, and JB wrote the initial draft of the manuscript. TM, YG, MP, ZH, WH, FW, and ES provided critical feedback and edited the manuscript.

